# Exploring the views of people with stroke regarding aerobic exercise participation during inpatient and outpatient rehabilitation: a qualitative descriptive study

**DOI:** 10.1101/2025.05.14.25327504

**Authors:** Azadeh Barzideh, Augustine Joshua Devasahayam, Ada Tang, Elizabeth L Inness, Susan Marzolini, Sarah Munce, Kathryn M Sibley, Avril Mansfield

## Abstract

**Objective:** We aimed to explore views of people with stroke regarding aerobic exercise participation during rehabilitation.

**Design:** Qualitative descriptive study informed by a pragmatic worldview.

**Setting and participants:** People with stroke attended online or in-person one-on-one semi-structured interviews focused on their general attitudes about aerobic exercise, and their capability, motivation and opportunities to exercise that have or have not been provided during rehabilitation after stroke.

**Data analysis:** Codebook thematic analysis was performed by two independent coders. Results: Thirteen people, 2 to 10 months post-stroke participated in the interviews. Six themes were identified: 1) having an exercise program routine and trusting the physiotherapist during rehabilitation facilitated doing exercise ; 2) emotions can make exercise during rehabilitation more or less difficult; 3) limited physical ability post stroke leads to poor exercise self-efficacy and sense of control; 4) knowledge of what exercise is and its benefits affects perseverance in exercising during rehabilitation; 5) personal identity affects perseverance in doing exercises during rehabilitation; and 6) environmental factors facilitate exercise performance (consisting of two sub-themes of supportive social environment promotes exercise participation, and more resources (e.g., time, space, staff, other programs) facilitate exercising during and after rehabilitation).

**Conclusions:** People with stroke are more likely to engage in aerobic exercise if it is incorporated into their treatment plan. This novel finding could help ensure physiotherapists prescribe aerobic exercise during stroke rehabilitation.

## Introduction

Functional improvement after stroke largely depends on participation in stroke rehabilitation, which may include physiotherapy, occupational therapy and speech-language pathology^1–3^. Participation in stroke rehabilitation is associated with reduced risk of mortality, death or dependency, need for institutionalization and length of hospital stay^4^.

Individually tailored aerobic training, aiming to enhance cardiovascular endurance, is a recognized component of stroke rehabilitation^5^. According to the recent Canadian Stroke Best Practice Recommendations, people with stroke, once medically stable, should participate in aerobic exercise at least 3 times per week for at least 8 weeks with a progressive increase in duration to a minimum of 20 minutes (excluding warm up and cool down)^5^. Including aerobic exercise as part of stroke rehabilitation provides the opportunity for people with stroke to receive support, counsel, and extrinsic motivation from physiotherapists; to exercise safely; and to build confidence and self-efficacy for exercising on their own after discharge.^6,7^ However, participation in aerobic exercise during rehabilitation is low^8,9^; less than 50% of eligible people with stroke participate in aerobic exercise during rehabilitation^8,9^.

Although aerobic exercise is an important component of stroke rehabilitation, only one study published in 2017 examined factors facilitating or impeding participation in aerobic exercise early in rehabilitation from patients’ perspective^10^. This quantitative study surveyed 33 patients at three Canadian rehabilitation hospitals to evaluate the self-efficacy and outcome expectancy of people with stroke from aerobic exercise^10^. This study found that people with stroke were willing to participate in aerobic exercise within the first week of inpatient rehabilitation^10^. In this study, inability to perform aerobic exercise, lack of social support from family, and lack of information were the reported barriers to aerobic exercise participation from the perspectives of people with stroke^10^. However, the role of other individual factors, such as motivation, fatigue, goals, and previous exercise experience, were not explored in this study. These factors are important to consider because evidence shows that tailoring interventions to each individual’s functional level and preferences improves uptake of and adherence to physical activity^11–13^. Furthermore, this quantitative survey study was unable to explain how or why things are happening, or to explain complex social or cultural phenomena^14^.

To tailor exercise interventions, individual factors such as psychosocial factors, symptoms, and physical activity history are assessed and then used to select the appropriate intervention components for each individual^15^. These components may include specific behaviour change techniques, parameters of intervention delivery, and exercise prescriptions to improve physical activity participation^16–19^. In this study, using qualitative methods, we aimed to explore patients’ perceptions of participation in aerobic exercise during rehabilitation. We also investigated the factors that facilitate or hinder participating in aerobic exercise during rehabilitation from the perspective of people with stroke.

## Methods

### Study design

We conducted a theoretically informed qualitative descriptive study guided by a pragmatic worldview^20,21^. The standards for reporting qualitative research (SRQR)^22^ were followed in reporting this study. The theoretical frameworks that were used in this study are the Theoretical Domains Framework (TDF^23^) and the Capability Opportunity Motivation Behaviour Model for Behaviour Change (COM-B^24^). TDF is a model for behaviour change that integrates 33 theories and 128 key theoretical constructs related to behaviour change and synthesises them into a single 14-domain framework^23,25^. This framework assesses implementation and other behavioural problems and informs intervention design. That is, it provides a theoretical lens to view the cognitive, affective, social and environmental influences on behaviour^25^. Although the TDF was originally developed to better understand the behaviours of healthcare professionals, it has been extended to other groups in which changing behaviour is important, including various patient groups^25^. The TDF informed creating the codebook and the deductive aspect of data analysis (e.g., primary TDF domains). The COM-B is a model for understanding behaviours with a goal of behaviour change^24^. COM-B was used to develop the interview questions (i.e., asking about previous and current physical capability of people with stroke, their motivation to exercise, and opportunities or lack thereof to exercise during rehabilitation). The behaviour change wheel maps appropriate behaviour change techniques to sources of behaviour (i.e., Capability, Opportunity and Motivation)^24^ and TDF domains are also mapped onto these sources of behaviour (i.e., COM-B segments)^26^.

### Participant selection and setting

People who were undergoing rehabilitation after stroke were eligible to participate in the study; participants were recruited from three urban rehabilitation hospitals in Ontario, Canada (Toronto Rehabilitation Institute – University Centre, Toronto Rehabilitation Institute – Rumsey Centre, and St. John’s Rehabilitation – Sunnybrook Health Sciences Centre). Two sites provide both in-patient and out-patient stroke rehabilitation, and one site offers out-patient rehabilitation only. The estimated number of patients with stroke admitted per year for each centre are: Toronto Rehabilitation Institute – University Centre: 450, Toronto Rehabilitation Institute – Rumsey Centre: 140, and St. John’s Rehabilitation – Sunnybrook Health Sciences Centre: 340. The research ethics boards of all participating sites approved the study. Participants were excluded if they had insufficient English language, communication and/or cognitive abilities to complete the interviews, as determined in consultation with the patients’ healthcare team. Eligible participants were identified by a member of the patients’ healthcare team, who asked the potential participants if they were interested in speaking with a research assistant regarding the study. If patients agreed, they were approached by the research assistant, who discussed the study with them, answered any questions that they may have had, and obtained written informed consent.

### Data collection

Data collection occurred via one-on-one semi-structured interviews with participants. The interview guide is provided in the supplementary material. These interviews were conducted by the PhD candidate (AB) via Microsoft Teams (Version 1.5.00.17656, Redmond, Washington, USA) or in person at the hospital if requested, from May 2021 to April 2023.

We aimed to conduct the interviews towards the end of participants’ rehabilitation stay, or soon after discharge from rehabilitation. The interviews began by providing a definition of aerobic exercise to participants based on Heart and Stroke Foundation of Canada’s lay definition; that is, any continuous activities or exercises that raise the heart rate or increase the breathing rate are considered aerobic exercise^27^. The following examples were then provided to the patients “for example, these exercises might include exercise on a piece of equipment like a stationary bike or stepper, walking either on the ground or on a treadmill”. Participants were then asked questions that started with their general attitudes about exercise and continued with their previous and current capability for exercise, motivation to exercise, and opportunities that have or have not been provided to them during rehabilitation.

The interview guide was pilot tested for feedback regarding clarity of the questions with one person with stroke who was discharged from rehabilitation more than two years previously. All interviews were audio-recorded and transcribed verbatim. The interviewer (AB) wrote field notes after each interview. Interviews lasted between 30 minutes to 1 hour.

### Sample size justification

We considered interpretive and pragmatic judgements to determine our sample size^28^. To make the interpretive judgement, we compared information power^29^ (as opposed to saturation) of our study to two recent studies with similar objectives and methodologies^30,31^. Our study was similar to these two studies on most elements of information power such as having a focused aim in the study and specificity of the participants’ experiences^29–31^.

However, for the latter study, the interviewers had more experience in the field compared to our study or the former study^31^. Therefore, we concluded that we needed to conduct more interviews in our study (i.e., more than ten); meaning our sample size should be closer to that in the Levy et al., study^30^ (i.e., in this study saturation was reached at eighteenth interview). Feasibility considerations (i.e., rate of refusal to participate in the study and resource constraints) led us to stop data collection after thirteen interviews. With these considerations, we believe we had a sufficient number of participants to contribute to knowledge in the field.

### Data trustworthiness

To enhance trustworthiness, we used the following methods: triangulation by the way of variation in data source and an investigator level triangulation^32,33^; generalizability by providing a rich description of the study sample^34^; and confirmability of the findings by using triangulation and leaving an audit trail between the raw data and interpretations^32,34^

### Data analysis

We used codebook thematic analysis^35^. A research assistant first transcribed each audio recording verbatim. Researcher AB listened to the audio recordings again and edited the transcripts for accuracy. Data analysis started concurrently with data collection following the six stages of thematic analysis^36^. After data transcription, two members of the team (AB and AM) read and familiarized themselves with the first three transcripts. To facilitate organization and analysis of qualitative data, the transcripts were entered into NVivo (V.12, QSR International, Burlington, Massachusetts, USA), which was also used for coding. TDF guided deductive coding. Concepts that could not be coded deductively were coded inductively. Two coders (AB and AM) with different backgrounds (physiotherapy and kinesiology) coded three transcripts separately. The coders then met to compare the labels and agreed on a set of codes to apply to all subsequent transcripts (i.e., a codebook was made). The codebook consisted of a table of the parent codes (i.e., the primary TDF domain), sub-codes, definitions of codes, and the example quotes. AB coded the remaining transcripts and met with AM to discuss any excerpts where she was uncertain. The final codebook, which did not change after coding the first 8 transcripts, was applied to all transcripts. AB subsequently drafted the themes that would explain the data, and the research team revised the themes until consensus was reached.

### Researcher characteristics and reflexivity

AB is a physiotherapist trained in Iran and Belgium. She had practice experience working in acute, sub-acute and chronic stroke care hospital units in both countries. Therefore, although she is familiar with stroke rehabilitation culture, not practicing in Canada helped her adopt an etic ontological position in this study. At the same time, being familiar with the stroke culture helped her during the interviews to probe the participants to better understand their experiences regarding participation in aerobic exercise. This helped providing in-depth accounts of people with stroke from aerobic exercise participation during rehabilitation in Canada.

AM, who was the second coder, is a female Scientist with a background in kinesiology and 12 years of experience as an independent researcher in the field of neurorehabilitation; she is the principal investigator in this study. AM has significant knowledge of exercise prescription post-stroke. She has conducted several studies of exercise post-stroke.

## Results

Forty-eight eligible people from 3 different urban rehabilitation centres were invited to participate in the interviews Of the 48 people invited, 35 declined and 13 consented to participate in the interview. The mean age of the sample was 64 (standard deviation=11.56) years. All participants were 2-10 months post-stroke at the time of the interview. Table 1 shows the characteristics of study participants. While the interviewer intended to elicit answers specific to aerobic exercise, respondents consistently referred to any exercise they did during rehabilitation. That is, most participants did not distinguish between the exercise types e.g., aerobic training, resistance training, functional exercises etc. Therefore, while the questions were specific to aerobic exercise, responses were directed toward any exercise that participants did during rehabilitation. Participants mostly talked about what made it more or less difficult for them to exercise as opposed to what completely prevented them from doing exercise. Participants reported continuing to exercise despite the challenges they faced during rehabilitation, and most were extremely satisfied with their experience with rehabilitation. All the names presented in this table and throughout the paper are pseudonyms.

**Table 1.** Participant characteristics. *Whether aerobic exercise was included in treatment plan (prescribed) and whether or not the people with stroke participated were based on their hospital charts.

| Patient | Sex | Age range | Admission | Stroke type | Affected brain side | Prescribed aerobic exercise* | Participated in aerobic exercise |
| --- | --- | --- | --- | --- | --- | --- | --- |
| Bart | Male | 60-70 | In-patient | Infarct | Left | No | No |
| Eric | Male | 50-60 | Out-patient | Infarct | Both sides | No | No |
| Kyle | Male | 40-50 | Out-patient | Haemorrhagic | Left | No | No |
| Marco | Male | 50-60 | Out-patient | Infarct | Left | No | No |
| Natalie | Female | 70-80 | Inpatient | Infarct | Left | No | No |
| Timothy | Male | 80-90 | In-patient | Infarct | Left | No | No |
| Gabriela | Female | 80-90 | In-patient | Infarct | Left | No | No |
| Curt | Male | 60-70 | Inpatient | Infarct | Right | No | Yes |
| Alicia | Female | 60-70 | In-patient | Infarct | Left | No | Yes |
| Corrine | Female | 70-80 | In-patient | Haemorrhagic | Right | Yes | Yes |
| Jed | Male | 40-50 | Out-patient | Infarct | Left | Yes | Yes |
| Maurice | Male | 50-60 | Out-patient | Lacunar | Left | Yes | Yes |
| Reed | Male | 70-80 | In-patient | Infarct | Left | Yes | Yes |

## Themes

### Overview of the themes

Overall, six themes regarding the factors that influence people with stroke participating in aerobic exercise during rehabilitation were identified: 1) having an exercise program routine and trusting the physiotherapist during rehabilitation facilitated doing exercise; 2) emotions can make exercise during rehabilitation more or less difficult; 3) limited physical ability post stroke leads to poor exercise self-efficacy and sense of control; 4) knowledge of what exercise is and its benefits affects perseverance in doing exercises during rehabilitation; 5) personal identity affects perseverance in doing exercises during rehabilitation; and 6) environmental factors facilitate exercise performance (consisting of two sub-themes of supportive social environment promotes exercise participation; and more resources (e.g., time, space, staff, other programs) facilitate exercising during and after rehabilitation). The inductive and deductive codes are presented in Table 2.

**Table 2.** Themes and their TDF constructs.

| Theme | TDF construct (domain)/<br>Inductive code | Findings |
| --- | --- | --- |
| Having an exercise program routine and trusting the physiotherapist during rehabilitation facilitated doing exercise | Inductive | All participants whose physiotherapist included aerobic exercise in their treatment plan reported that they participated, whether or not they found it difficult. No participants declined participating in aerobic exercise during their stroke rehabilitation. |
| Emotions can make exercise during rehabilitation more or less difficult | Anxiety<br>Fear<br>Depression<br>Positive/negative affect (Emotions) | Participants experienced a wide range of positive and negative emotions, which influenced the level of difficulty they perceived while exercising during rehabilitation. Managing emotions seemed important for patients. |
|  | Empowerment<br>(Beliefs about Capabilities) | Some patients felt empowered when they saw other patients progress, or when they observed their own progress. |
| Limited physical ability post stroke leads to poor exercise self-efficacy and sense of control. | Self-efficacy<br>Perceived<br>-behavioural control<br>(Beliefs about Capabilities) | Stroke negatively influenced participants' self-confidence to exercise during and soon after rehabilitation. Many of the participants wanted control over their behaviours, but they felt that they could not always have this control for safety reasons. |
|  | Ability (Skills) | Most people felt they did not have the ability to exercise (e.g., felt weak and fatigued), especially earlier post-stroke. |

TDF= Theoretical domain framework

Themes and subthemes
| Theme | TDF construct (domain)/<br>Inductive code | Result |
| --- | --- | --- |
| Knowledge of what exercise is, and its benefits affects perseverance in doing exercises during rehabilitation | Knowledge (Knowledge) | Participants with pre-stroke exercise experience persevered with participation in exercise during rehabilitation. |
| Personal identity affects perseverance in doing exercises during rehabilitation. | Identity (Social/Professional Role and Identity) | The people who identified as being independent and wanting to do everything themselves/not giving up easily/not waiting for other's help persevered in doing exercise too. |
| Environmental factors facilitate exercise performance (two subthemes of supportive social environment and more resources) | Social support (Social influences) | Having encouraging and supportive physiotherapists during rehabilitation helped patients feel supported and helped them to exercise during rehabilitation. |
|  | Material resources (Environmental Context and Resources) | Some patients preferred longer duration of rehabilitation, more space in the gym, and having stretching and massage therapy. |

#### 1) Having an exercise program routine and trusting the physiotherapist during rehabilitation facilitated doing exercise

Participants reported that they performed any exercise that their physiotherapists incorporated in their treatment plans (including aerobic exercise), even if they perceived it difficult or fatiguing; no participant in our study reported declining participating in any exercise that was offered to them. Most participants did not distinguish between various types of exercise they completed during rehabilitation. Most participants said that they trusted their physiotherapists to do what was necessary for them. Curt explained,

> I trusted them [physiotherapists] to allow me to do as much as they thought was appropriate.

In response to the question “what were the factors that helped you to exercise during rehabilitation when you were in in-patient rehabilitation?” Jed responded:

> I think what really helps is that, I know it sounds kind of odd and obvious, but if they give you the chance to do it, then it is great, that is what helps. [It] is the opportunity, you know, or them suggesting or taking the initiative to have you try something. That, that is, that in and of itself is helpful.

One participant, whose stroke did not affect their physical function, only received occupational therapy, not physiotherapy, during rehabilitation; however, they mentioned that they would have liked to receive professional advice about how to exercise, how much to do, with what intensity, and what to do to relieve the pain caused by exercise.

Another participant found it difficult to continue to exercise after discharge from rehabilitation when they no longer had a structured exercise program and routine. They believed having a routine facilitates doing exercise during rehabilitation. Another participant was surprised that there was no therapy on weekends.

#### 2) Emotions can make exercise during rehabilitation more or less difficult

Participants experienced a wide range of positive and negative emotions, which influenced the level of difficulty they perceived while exercising during rehabilitation. Negative feelings included having a low mood, feeling shocked, disappointment because their progression was not as fast as they would like, feeling that stroke was catastrophic, and feeling that exercises were at times tiring and frustrating. Bart elaborated:

> it’s a big scare at the beginning……[I felt] I’m useless, I can’t do this, I can’t walk, I can’t pick that up, I can’t hold that, I can’t carry a plate, I can’t carry a cup of water and stuff and it was really depressing at the beginning.

One participant sometimes felt overwhelmed during inpatient rehabilitation, so they went outside and got fresh air and talked to other patients; this helped them to manage their feelings and enabled participation in exercise the next day. Three patients reported feeling bored during in-patient rehabilitation for various reasons, including not having much to do outside formal therapy time, not having music/TV while doing aerobic exercise on the equipment, and upper limb exercises being mundane, monotonous and repetitive. Many participants expressed feeling depressed after the stroke because they could not do what they used to be able to do, because they felt very tired, or because of being in the hospital environment.

Positive emotions made it easier for participants to exercise, including feeling accomplished, satisfied with their rehabilitation process, lucky or impressed with their therapist and that they could start rehabilitation right away, encouraged by their healthcare team, and ‘runner’s high’ after the exercise. Feeling motivated and empowered also positively affected performing exercise during rehabilitation. Almost all patients believed that rehabilitation/exercise would get them as close as possible to where they were before the stroke, or would put them in the mind frame of one day getting there. Many believed that doing any exercise would help recovery, whereas some believed if they did not exercise, their mobility will decline. Many patients believed sitting in the bedroom all day during rehabilitation is harmful, so they reported exercising (mostly in bed/standing) or going for walks outside scheduled physiotherapy sessions on their own. These beliefs motivated patients to exercise during rehabilitation, even if it was difficult for them. A few patients were motivated to exercise more during rehabilitation, and some were specifically talking about aerobic exercise.

In response to the question “what helped you to exercise during rehabilitation?”, Alicia said,

> I think it’s my eagerness to get well again. And because [when] I was at that rehabilitation centre, I realized it’s always full house, getting a spot to start my exercise is, I was so lucky. So I keep telling myself, you know, take advantage of all the professionals here and then and get well as soon as possible. That’s why I do all my exercise.

Other incentives to exercise included observing their own progress, help and encouragement from their therapists, and challenges therapists set for patients. One participant’s motivation came from their physiotherapist’s and other people’s encouragement, and seeing videos that their physiotherapists showed them about improvement post-stroke.

Some participants felt empowered when they saw other patients progress, or when they observed their own progress. Feeling empowered led to trying to be more physically active and being more determined to exercise during rehabilitation, showing adaptive behaviour (e.g., finding ways to function with the use of less affected arm), putting more effort to get independent and receiving the care they wanted. One participant (during out-patient rehabilitation) asked the physiotherapists to prescribe aerobic exercise to them because they wanted to try the equipment and buy one at home. When physiotherapists accommodated participants’ requests (e.g., changing the time of the physiotherapy session) and offered individualized exercises based on participants’ goals, they empowered participants to exercise. One participant believed that having more in-room exercises would help them build the habit of exercising on their own.

#### 3) Limited physical ability post stroke leads to poor exercise self-efficacy and sense of control

Some participants said they had the ability to do more during rehabilitation, but most felt they did not have the ability to exercise (e.g., because they felt weak and fatigued), especially earlier post-stroke. Some participants believed that they were still not where they wanted to be in terms of their physical abilities after discharge. Most participants reported feeling fatigue after their strokes, which in some cases affected their ability to exercise. Weakness and fatigue led two participants not to exercise after rehabilitation. One participant felt exercise was always an effort during rehabilitation due to weakness. Feeling tired after performing activities of daily life such as grocery shopping affected Eric’s ability in doing home exercises that his outpatient physiotherapist prescribed to him. He elaborated,

> So I think that [grocery shopping and carrying the bags] works for, in place of the exercises that they give me here like pictures or everything and they really are good exercise, I know that. Everyone, all the physical therapists have showed me what to do. I know they’re good exercises because I’ve done a lot of them with occupational therapists, physiotherapists, here in the last month. But when I, at home, like I’m just too tired, like to go, to do groceries and that that’s as much as I could do in one day.

Stroke negatively influenced participants’ self-confidence to exercise during and soon after rehabilitation. That is, stroke-related disabilities impacted their ability to exercise, which in turn impacted participants’ confidence. However, participants gradually got their confidence back when they saw improvements. Lack of confidence led to perceived difficulty in exercising during rehabilitation, and no or low intensity exercise after rehabilitation, while gaining confidence back helped patients to exercise with more ease. In response to the question: “how did your stroke affect you physically?”, Timothy said:

> [Stroke] took away a lot of my confidence. It took away a lot of my confidence. There’s a certain level of fear, for my independence is gone. Well, not gone. But my independence has been diminished.

Many participants wanted control over their behaviours and their situation after stroke because they wanted to get better, but they felt that they could not always have this control for safety reasons. Three patients specifically mentioned that they would have liked to exercise after formal physiotherapy time e.g., getting on the equipment or doing some other activities that do not need supervision or only with the supervision of physiotherapy assistants. However, they further stated that using exercise equipment outside formal therapy sessions was ‘not allowed’. Some patients stated they were ‘not allowed’ to do any activity without a nurse or a physiotherapist. One patient believed that physiotherapists ‘play it safe’ during rehabilitation, so it was important to them to communicate with the physiotherapist about what they can and cannot do (based on what their body can/cannot do). Maurice elaborated,

> Actually usually those people will not give you all the things that you can do. Yeah, they usually play safe, and don’t give all the things that normal people can do… I think they should allow people to do things to their capacity.

#### 4) Knowledge of what exercise is and its benefits affects perseverance in doing exercises during rehabilitation

Participants that previously exercised before their stroke persevered with participating in exercise after their stroke. Some participants believed they have good knowledge of exercise, but most believed they did not have enough knowledge. Participants with good knowledge (e.g., knew various exercise types and their benefits) had the intention to or were already exercising after discharge from rehabilitation. Participants who did not have enough knowledge still tried to keep active, but they mostly performed physical activities rather than structured exercise after discharge from rehabilitation. One participant said they had not received education on the stroke trajectory, what rehabilitation is, and how to use aerobic exercise equipment from their physiotherapist.

#### 5) Personal identity affects perseverance in doing exercises during rehabilitation

Participants who identified with being an independent person and wanting to do everything themselves/not giving up easily/not waiting for other’s help persevered in doing exercise. However, one participant said they were lazy but still exercised during rehabilitation, and another did not like exercise but exercised during rehabilitation as they were motivated to leave the rehabilitation facility. One participant believed they were very susceptible and open to new exercises and new initiatives from physiotherapists and wanted to do different activities during rehabilitation. Another participant said, “I’m a bit of a chicken”, which led to them being very careful and very cautious and not to push much. Gabriella in response to the question “what helped you to exercise during rehabilitation?” said,

> Probably saying to myself “you can do this”, you know “you’ve never let things challenge you before”, I’ve always been that kind of a person.

#### 6) Environmental factors facilitate exercise performance

##### Supportive social environment promotes exercise participation

Having encouraging and supportive physiotherapists during rehabilitation helped participants feel supported and helped them to exercise during rehabilitation. Additionally, while not the focus of the study, many participants also talked about having encouraging and supportive people around after discharge from rehabilitation. Participants reported that having supportive family members and friends helped them continue exercising after rehabilitation, whereas not having support or encouragement from physiotherapists or family/friends led to being less active outside formal therapy time during and after rehabilitation, respectively. Eric, who did not have family support to help with activities of daily living such as grocery shopping, cooking etc., could not perform home exercises during outpatient rehabilitation as he did not have the energy and felt fatigued after completing these activities himself. This experience affected the advice they would give to other people with stroke:

> It [patient’s advice] would depend if they [other people with stroke] have someone to look after them like a family unit or they’re on their own. It’s different pieces of advice. If they had a family looking after them, I would say do everything that you’re told, do everything that they show you in the rehabilitation, yeah. Because it’s very good exercise. If it was someone that has no one to look after them, it’s a totally different uh recommendation. I would just say: listen to them, take the pictures (i.e., pictures of home exercises) home and keep it for later and just do the best you can. Because you have to do everything yourself and that’s a big difference for one to have.

##### More resources (e.g., time, space, staff, other programs) can facilitate exercising during and after rehabilitation

One participant said they would increase the length of their inpatient rehabilitation if they could because they were not ready to be discharged, and this negatively affected their ability to exercise afterward. Some participants believed the length of physiotherapy sessions could have been longer. Some participants said more space in the gym would be better and some believed more physiotherapists are needed. Another participant believed stretching and massage therapy should be added to rehabilitation to ease muscle tightness. Additionally, while not the focus of the study, many participants also talked about having access to exercise plans or devices after discharge from rehabilitation. Participants reported that physiotherapists generally advised them to continue to exercise post discharge and provided them with home exercise plans (based on participants’ access to resources), which helped them know what to do after discharge.

## Discussion

We explored the perspectives of people with stroke on factors that facilitate or impede participation in aerobic exercise during rehabilitation. Most participants were not concerned about the exercise types they were prescribed and did not distinguish between various exercise types. They performed any exercise that their physiotherapist incorporated in their treatment plans (including aerobic exercise), even if they perceived it difficult or fatiguing. These findings suggested that one of the important contributors to participation in aerobic exercise during rehabilitation after stroke is whether the physiotherapists include it in the patients’ treatment plans.

Patients’ emotions affected the difficulty people with stroke perceived while exercising during rehabilitation. However, these factors did not limit participation of people with stroke in exercise. The association between emotions and functional gains and physical abilities of people with stroke has been previously reported^37^. Having negative emotions (e.g., feelings of distress, upset and being scared) affected knee muscle strength and gait quality variables (e.g., velocity, cadence), and independence in performing activities of daily life negatively^38^. These findings emphasize that physical and emotional recovery of people with stroke are connected^39^. Having emotional responses after stroke was also reported by Last et al., where the emotional responses triggered by new challenges and living with a changed body appeared to negatively impact patients’ desire to participate in rehabilitation in some cases^40^.

Although people with stroke experience depression, another study has also showed that most people with stroke during rehabilitation completed rehabilitation activities regardless of motivation,^41^ similar to our findings. This may reflect the influence of having an exercise program and encouraging physiotherapists during rehabilitation. Physiotherapists’ contribution to the patient empowerment process is crucial, and physiotherapists should be aware their own role in motivating and empowering people with stroke. We observed that feeling empowered promoted physical activity/exercise outside therapy time during rehabilitation. A recent qualitative study exploring the lived experience of empowering people with stroke during rehabilitation supports this notion and identifies seven themes^42^. Three of these seven themes focus on the role of physiotherapist in patient empowerment^42^.

Rehabilitation professionals tend to assume people with stroke who understand the purpose of rehabilitation and what is expected of them are highly motivated^43^. That is, patients who are more likely to understand the nature and purpose of rehabilitation are considered as highly motivated by rehabilitation professionals^43^. Therefore, providing patients with information about their stroke, the rehabilitation processes, and purpose of rehabilitation may increase motivation in people with stroke^43^ although knowledge alone may not be sufficient to change the behaviour^44^. As our study showed, participants who exercised before stroke, and therefore knew various exercise types and their benefits, persevered with participating in exercising during rehabilitation. Gaining knowledge about rehabilitation empowers people with stroke and diminishes their feeling of powerlessness^45^.

Only one other study has previously explored views of people with stroke regarding aerobic exercise during rehabilitation^10^. However, a few studies have explored the factors that influence participation in stroke rehabilitation in general and have reported findings similar to ours^40,46,47^. For example, Last et al, explored the perspectives of people with stroke on barriers and facilitators to participation in hospital-based stroke rehabilitation, and identified four themes^40^: environmental factors (i.e., physical and social environments, resources); components of therapy (including patient-therapist interactions, amount of therapy, and personalized rehabilitation); physical and emotional well-being (including fatigue, and emotional adjustments); and personal motivators (including resuming life roles, and attitudes towards rehabilitation)^40^. Similar to our findings, Last et al., reported that when interventions include activities that are meaningful to patients and help patients to resume their valued life roles, it empowers them to participate in those activities^40^. In our study, almost all participants said that they were very satisfied with their physiotherapists and the rehabilitation process in general and only one patient said they did not receive education on what rehabilitation is and how they should use the equipment.

Feeling fatigued was reported by most of our participants, however, only two participants said fatigue affected their ability to exercise and limited the amount of exercise they can do at home. One of these participants reported fatigue when performing activities of daily living such as grocery shopping and the other when doing exercise at home. Feeling fatigued after doing activities of daily living upon reintegration into everyday life after discharge from rehabilitation has been reported among people with neurological disabilities although the effects of fatigue on performing exercise during or after rehabilitation were not reported^48^. Aerobic exercise has been shown to improve cardiovascular fitness and build up the capacity to participate in strenuous activities without fatigue^49^. In our study, physical activity was valued by participants, and they tried to stay active (e.g., walking in the corridors) outside formal therapy time as much as possible. While some studies have suggested that people with stroke spend most of their time in their room, being solitary and inactive^50,51^, others have supported our finding that people with stroke try to be active outside of therapy time^52,53^. There is preliminary evidence suggesting that improving the ease of navigation of hospital environments, with attractive and accessible communal areas and fewer single-bed patient rooms, may increase activity levels of people with stroke during inpatient rehabilitation^54^.

We intended to perform stratified purposeful sampling to select study participants (i.e., based on site and participation in aerobic exercise) and ensure that a variety of participants and perspectives were captured^55^. However, many people with stroke refused to participate in the interviews for various reasons (e.g., being overwhelmed with healthcare appointments). Therefore, we recruited and interviewed any participant who consented to the interview. We were still able to capture a variety of experiences (e.g., varied living situations and attitudes towards exercise not being prescribed physiotherapy during rehabilitation) from different urban rehabilitation sites and people of different age ranges. Unfortunately, most of the participants in this study did not have aerobic exercise included in their treatment plans during rehabilitation.

This study explored perspectives of people with stroke of aerobic exercise participation during rehabilitation and helped us to obtain a better understanding of their challenges. Our participants reported participating in any exercise (including aerobic exercise) that is included in their treatment, even if they feel it is challenging for them; meaning that people with stroke participate in aerobic exercise if it is included in their treatment plan. This novel finding could help physiotherapists with their clinical decision-making during rehabilitation of people with stroke.

## Clinical messages

- Emotions, limited physical ability post stroke, and environmental factors can make exercise feel more or less difficult during rehabilitation. But people with stroke participate in aerobic exercise if it is included in their treatment plans regardless of how challenging it is.
- Personal identity of people with stroke together with their knowledge of what exercise is and its benefits affect perseverance in doing exercises during rehabilitation.
- Having an exercise program routine and trusting the physiotherapist during rehabilitation facilitate doing exercise.

## Supporting information

Interview guide for patients

## Data Availability

Consent was not obtained for interview transcripts to be shared with third parties.

## Acknowledgments

Authors would like to thank Kay-Ann Allen and David Jagroop for their assistance with participant recruiting, and Devanshi Joshi, Saira Thavaneethan, and Sarah Thompson for their assistance during the interviews and for transcribing the interviews.

