## Supplementary material for "Exploring the views of people with stroke regarding aerobic exercise participation during inpatient and outpatient rehabilitation: a qualitative descriptive study": Interview guide for patients

Supplemental material

Interview guide for patients

*Interviewer will first remind participants about the purpose of the study, confidentiality, and consent to be audio recorded before proceeding with the questions below.*

*COM-B items probed with individual questions: C=Capability, M=Motivation, O=Opportunity, B=Behaviour*

I am going to ask you questions about your experiences with aerobic exercise during your stroke rehabilitation. Aerobic exercise includes “any continuous activities or exercises that raise your heart rate or increase your breathing rate”. For example, these exercises might include exercise on a piece of equipment like a stationary bike or stepper, walking either on the ground or on a treadmill. You are free to skip any questions that you do not want to answer, and you can end the interview at any time.

Are you ready to begin?

1. Tell me about some activities you have done, either before or after your stroke, that could be considered aerobic exercise. [**C**,**B**]

*Prompts*

What kinds of activities did you do? How often did you do them? How did they affect your heart rate/breathing rate? [**C**,**B**]

Did/do you enjoy this type of activity? What did you like about it? What did you not like about it? [**C**,**M**]

2. Can you tell me a bit about how your stroke has affected you? [**C**]
3. Can you tell me about your experiences with rehabilitation after your stroke?
4. What have your experiences been with doing aerobic exercise during rehabilitation for your stroke?

*Prompts*

Did your physiotherapist prescribe aerobic exercise during your rehabilitation stay? [**O**]

*If physiotherapist prescribed exercise:* did you agree to participate in aerobic exercise? Why/why not? [**B**]

*If they did aerobic exercise:* What did you do? How often did you do it? How would you describe the level of challenge (e.g., too easy, too difficult, just right)? What was your understanding of the purpose of participating in aerobic exercise? [**C**,**B**]

How do you think you benefited from this type of exercise, if at all?

What was the hardest part about exercising? What was the easiest? [**C**,**M**]

*If physiotherapists didn’t prescribe exercise*: would you have liked to do aerobic exercise during rehabilitation? Why? [**M**,**O**]

Did you try to do aerobic exercise on your own outside of physiotherapy? What were your experiences with this? [**O**,**B**]

5. What are some of the challenges you’ve experienced with aerobic exercise during rehabilitation?
6. What things have helped you to exercise during rehabilitation?

*Prompts:*

Did you routinely exercise before your stroke (i.e., habits)? How did your experiences with exercise before your stroke influence your attitudes towards exercise after stroke? [**C**,**M**,**B**]

How would you describe your skill with/knowledge of how to exercise? [**C**]

Do you think aerobic exercise is important for people early after stroke? [**M**]

Was there anything in the hospital that helped or prevented you from participating in aerobic exercise? [**O**] Did your physiotherapist prescribe exercise for you? [Physiotherapist **B**, Patient **O**] Did your physiotherapist encourage you to exercise? [Physiotherapist **B** influence on Patient **M**]

Did your spouse, family members, of friends encourage or discourage you from exercising after your stroke? [**M**,**O**]

Were there any other factors that influenced your ability to participate in exercise during your rehabilitation program? [**C**,**M**,**O**]

*Depending on responses to question 2*: You told me before that your stroke affected your	; how does this impact your ability to participate in exercise? [**C**]

*Consider promoting about specific challenges; e.g., time/feeling busy, fatigue, depression/mood/mental health*

7. If you had the power to change something in rehab unit/service to make it easier for people with stroke to participate in exercise, what would you change? [**O**]

*Prompt*

Is there something that your physiotherapist or other member of your care team could have done to help you participate in exercise? [**O**]

Is there something that could have been done differently during your rehabilitation to make it easier for you to exercise? [**O**]

8. What would you tell another patient who is about to start an aerobic exercise program during stroke rehabilitation?

*Definition of aerobic exercise from Prout et al., 2017*

*Questions modified from Pak et al., 2015 and Prout et al., 2017*
